# Why do different people with Spinal Cord Injury have differing severity of symptoms with Autonomic Dysreflexia? Exploring relationships of vascular alpha-1 adrenoreceptor and baroreflex sensitivity after SCI

**DOI:** 10.1101/2024.05.02.24306772

**Authors:** Jennifer Dens Higano, Kathryn Burns, Geoffrey Smith, Ryan Solinsky

**Affiliations:** Mayo Clinic; Spaulding Rehabilitation Hospital

## Abstract

**Introduction:** Individuals with spinal cord injury (SCI) commonly have autonomic dysreflexia (AD) with increased sympathetic activity. After SCI, individuals have decreased baroreflex sensitivity and increased vascular responsiveness.

**Objective:** To evalate relationship between baroreflex and blood vessel sensitivity with autonomic dysreflexia symptoms.

**Design:** Case control

**Setting:** Tertiary academic center

**Patients:** 14 individuals with SCI, 17 matched uninjured controls

**Interventions:** All participants quantified AD symptoms using the Autonomic Dysfunction Following SCI (ADFSCI)-AD survey. Participants received three intravenous phenylephrine boluses, reproducibly increasing systolic blood pressure (SBP) 15-40 mmHg. Continuous heart rate (R-R interval, ECG), beat-to-beat blood pressures (finapres), and popliteal artery flow velocity were recorded. Vascular responsiveness (α1 adrenoreceptor sensitivity) and heart rate responsiveness to increased SBP (baroreflex sensitivity) were calculated.

**Main outcome measures:** Baroreflex sensitivity after increased SBP; Vascular responsiveness through quantified mean arterial pressure (MAP) 2-minute area under the curve and change in vascular resistance.

**Results:** SCI and control cohorts were well-matched with mean age 31.9 and 29.6 years (p=0.41), 21.4% and 17.6% female respectively. Baseline MAP (p=0.83) and R-R interval (p=0.39) were similar. ADFSCI-AD scores were higher following SCI (27.9+/-22.9 vs 4.2+/-2.9 in controls, p=0.002).

To quantify SBP response, MAP area under the curve was normalized to dose/bodyweight. Individuals with SCI had significantly larger responses (0.26+/-0.19 mmHg*s/kg*ug) than controls (0.06+/-0.06 mmHg*s/kg*ug, p=0.002). Similarly, leg vascular resistance increased after SCI (24% vs 6% to a normalized dose, p=0.007). Baroreflex sensitivity was significantly lower after SCI (15.0+/-8.3 vs 23.7+/-9.3 ms/mmHg, p=0.01). ADFSCI-AD subscore had no meaningful correlation with vascular responsiveness (R^2^=0.008) or baroreflex sensitivity (R^2^=0.092) after SCI.

**Conclusions:** While this confirms smaller previous studies suggesting increased α1 adrenoreceptor sensitivity and lower baroreflex sensitivity in individuals with SCI, these differences lacked correlation to increased symptoms of AD. Further research into physiologic mechanisms to explain why some individuals with SCI develop symptoms is needed.

## INTRODUCTION

Individuals with spinal cord injury (SCI) commonly have autonomic dysreflexia (AD),^1^ defined as an increase in systolic blood pressure (SBP) of greater than 20 mmHg from baseline.^2^ At times this may escalate to dangerous hypertension, and multiple studies indicate that individuals at risk have an average of over ten episodes of dysreflexia a day.^3,4^ Common symptoms of AD are flushing, diaphoresis, headache, piloerection, palpitations, blurred vision, and anxiety.^1,5^ However, much of AD can be asymptomatic or “silent”.^6,7^ When symptoms do occur, they are often variable among individuals, clouding the clinical presentation^8^ and making clear diagnostics elusive. One individual may reliably present with AD with fast onset headache and sweating when in need of repositioning, while another may only have mild tingling on their neck, signaling to them that it is time to empty their bladder. But why do people with SCI present so differently with autonomic dysreflexia?

Pathophysiologically, AD is characterized by an amplified sympathetic response to noxious stimuli below the level of SCI. This, in turn, leads to vasoconstriction and associated hypertension. In an attempt to compensate, vasodilation may occur above the level of injury, where autonomic control is intact. The increase in SBP from below level vasoconstriction is sensed at the carotid and aortic baroreceptors, triggering brainstem and vagal pathways to directly slow the heart.

The efficacy of this compensatory feedback loop is termed baroreflex sensitivity and serves as a quantifiable marker of compensatory ability (ms of slowing in R-R interval per mmHg increase in SBP).^9^ After SCI, baroreflex sensitivity has been shown to be generally decreased, though some individuals appear near normal.^10^ Decreases in baroreflex sensitivity have been postulated to be due to accelerated atherosclerosis^11^ and reversal of normal blood pressure feedback mechanisms^12^ – both conditions which may be variable depending on autonomic level and completeness of injury.

Clinically, this equates to variability in the heart rate responses of individuals with SCI when they have AD.^13^ As individuals often consciously perceive profound changes in heart rate, we reasoned that baroreflex sensitivity may in turn contribute to the differences in perceived symptoms of AD.

Similarly, another variable pathophysiologic feature of AD after SCI is arterial responsiveness. Sympathetically mediated vasoconstriction occurs in AD when released norepinephrine binds to α1 adrenoreceptors in the blood vessels. In some cases, decreased baseline levels of plasma norepinephrine are seen after SCI.^14-16^ These low baseline levels result in increased expression of α1 adrenoreceptors,^17^ making vessels potentially hyper-responsive to sympathetic activation.^18^ During AD, with unregulated sympathetic activity prompting release of these agonists, the vessel hypersensitivity could result in greater or more rapid vasoconstriction that could be related to increased AD symptoms.

Thus, we set out to identify relationships between symptoms of AD and variable baroreflex and α1 adrenoreceptor sensitivities. We hypothesized that individuals with SCI would have increased, but variable, vessel α1 adrenergic sensitivity and lower baroreflex sensitivity when compared to uninjured controls, and that more extreme pathophysiology after SCI would be correlated with increased symptom severity of AD.

## METHODS

Following IRB approval, both individuals with SCI and matched uninjured controls were enrolled in this prospective case/control study. Participants with SCI were included if they had history of traumatic SCI with American Spinal Injury Association Impairment Scale (AIS) A-D injuries and level of injury from C1-T12,^19^ and were between 18-50 years old. These wide inclusion criteria (with individuals with AIS D SCI and those with levels of injury below T6 included) were preselected to encompass the breadth of those who have been documented to have AD after SCI.^20^ Exclusion criteria for all participants included current pregnancy, other non-SCI neurological disorder, history of clinically-diagnosed cardiovascular disease, hypertension, or diabetes. For uninjured controls, participants were excluded with a body mass index (BMI) greater than 30 kg/m^2^ to limit participants with undiagnosed cardiovascular disease and diabetes which may confound results.

To minimize diurnal variation in sympathetic activity, participants presented to our lab between 8AM and 10AM, at least 12 hours postprandial, at least 24 hours since refraining from consumption of caffeine or alcohol, and at least 48 hours after last exercising.^21,22^ Demographic characteristics and risk factors for cardiovascular disease (smoking history, antihypertensive/statin/aspirin treatment, BMI, race) were screened for all participants. Symptoms of AD were assessed with the Autonomic Dysfunction Following SCI survey (ADFSCI) for all participants, which is validated to correlate with the number of episodes of dysreflexia per day as tested with ambulatory blood pressure monitoring.^3,4^ We included uninjured controls as part of this data collection since although symptoms (headache for example) may be attributed to AD, symptoms may also occur in those without SCI as part of normal variability. Individuals with SCI further had verification of neurological level of injury and clinical completeness using the ISNSCSCI evaluation.^19^

### Investigational protocol

Participants rested supine for at least 15 minutes to equilibrate prior to the experimental protocol, which encompassed a battery of autonomic tests (NCT04493372). Participants were instrumented with a 5-lead ECG to derive heart rates/R-R wave intervals. Continual beat-to-beat blood pressures were recorded using a Finapres Blood Pressure System along with oscillometric blood pressure recording system for calibration. Beat-by-beat calf blood flow was recorded using a 4 MHz Doppler probe placed against the skin behind the knee to isolate the popliteal artery proximal to bifurcation and to derive flow velocities as an estimate of whole-limb blood flow. An IV was placed in the antecubital fossa for serial bolus phenylephrine infusions.

With instrumentation in place, bolus phenylephrine was serially administered using the Oxford technique to pharmacologically increase SBP by a goal of 15-40 mmHg (2.5 μg/kg to a max of 175 μg of phenylephrine). This technique has been safely performed since the 1960s in a wide range of individuals, including those with a recent myocardial infarction.^23^ Three trials were completed to ensure a reliable measurement with at least eight minutes (approximately two medication half-lives)^24^ allowed between boluses to limit carryover effects. Increase in SBP following bolus phenylephrine and accompanying change in cardiac slowing (by R-R interval change) were used to measure baroreflex sensitivity. By evaluating the magnitude of change in SBP and increase in vascular resistance, the protocol also allowed quantification and comparison of α1 adrenoreceptor sensitivity. All data was continuously recorded through the completion of the investigational protocol using LabChart and digitized for offline analysis.

### Data analysis

The AD subscore of the ADFSCI questionnaire was calculated and descriptive statistics were completed. For baroreflex sensitivity, baseline mean SBP and R-R interval were calculated for 30 seconds prior to bolus infusion. Changes in SBP and R-R interval from this baseline were calculated at the initial peak response of bolus phenylephrine and used to calculate baroreflex sensitivity (Figure 1). Per convention, increases in SBP less than 10 mmHg were not included due to insufficient baroreceptor stimulation and associated inaccurate gains.

**Figure 1:**
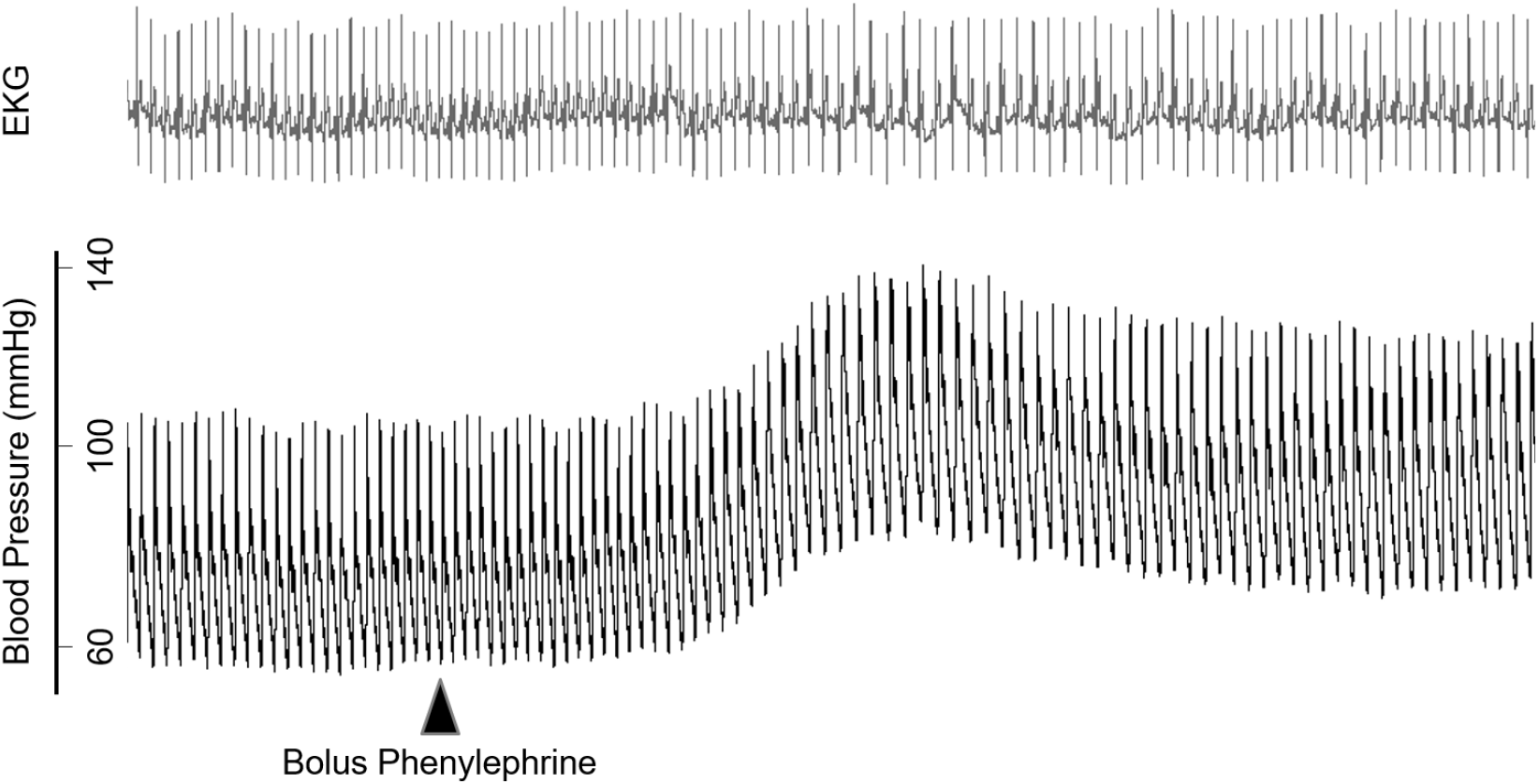
Example of heart rate and blood pressure responses to bolus intravenous phenylephrine.

To characterize α1 adrenergic receptor sensitivity, baseline mean arterial pressure (MAP) was calculated from a three-minute quiet supine resting period prior to each bolus. MAP area under the curve for the first two minutes from onset of the phenylephrine response was calculated relative to baseline. Baseline lower extremity vascular resistance was calculated from MAP and flow velocities. To further quantify the response, we evaluated changes in this vascular resistance following bolus phenylephrine. After confirming baseline differences between cohorts based on past literature,^25^ percent change was calculated. All measurements were normalized per μg of phenylephrine and kg of body weight, and the mean of the three boluses was used to calculate correlations to symptoms of AD via linear regression analysis. Statistical significance was set at a p-value of less than 0.05. Values are reported as means ± standard deviations.

## RESULTS

Thirty-one people were enrolled in this study, with the 14 individuals in the SCI cohort well matched by demographics and resting vitals to 17 individuals in the uninjured control cohort (Table 1).

**Table 1:** Study Cohort Demographics and Baseline Measurements. BMI= Body mass index; NLI= Neurological Level of Injury; AIS= American Spinal Injury Association Impairment Scale; MAP= Mean Arterial Pressure; SBP= Systolic Blood Pressure; DBP= Diastolic Blood Pressure; bpm= beats per minute; ADFSCI-AD= Autonomic Dysfunction Following SCI survey, Autonomic Dysreflexia subscore. Racial demographics for each cohort appear in Supplemental Table 1.

|  | Controls<br>(n=17) | SCI<br>(n=14) | p-value |
| --- | --- | --- | --- |
| Mean Age (range) | 29 (22-47) years | 32 (22-44) years | 0.41 |
| % Female | 18% | 21% | 0.80 |
| Mean BMI (range) | 23 (19-27) kg/m<sup>2</sup> | 23 (15-39) kg/m<sup>2</sup> | 0.91 |
| NLI C1-C8 | - | 7 |  |
| T1-T6 | - | 2 |  |
| T7-T12 | - | 5 |  |
| AIS A | - | 9 |  |
| B | - | 2 |  |
| C | - | 2 |  |
| D | - | 1 |  |
| Years since injury | - | 8.3 (1.9-38.7) |  |
| **Baseline measurements** |  |  |  |
| Mean resting SBP (SD) | 121.9 (11.6) mmHg | 118.9 (14.7) mmHg | 0.54 |
| Mean resting DBP (SD) | 67.9 (10.0) mmHg | 75.2 (14.4) mmHg | 0.13 |
| Mean resting heart rate (SD) | 60 (9) bpm | 61 (20) bpm | 0.25 |
| ADFSCI- AD mean (SD) | 4.2 (2.9) | 27.9 (22.9) | 0.002 |

As expected, the SCI cohort had significantly lower average lower extremity vascular resistance (4.3 ± 1.4 U) compared with controls (8.3 ± 5.2 U, p= 0.007) at baseline. Given this baseline difference, percent change in vascular resistance was confirmed to be statistically appropriate. Baroreflex sensitivity was calculated and found to be significantly lower after SCI compared to controls (15.0 ± 8.3 ms/mmHg for SCI cohort vs 23.7 ± 9.3 ms/mmHg for controls, p=0.01, Figure 2A). Individuals with SCI had significantly larger MAP increases to a normalized dose of IV phenylephrine, with 2-minute AUC analysis showing an over four times larger average blood pressure response (0.26 ± 0.19 mmHg·s/kg·μg) compared to controls (0.06 ± 0.06 mmHg·s/kg·μg, p=0.002, Figure 2B). Matching these findings, mean lower extremity vascular resistance increased to a greater degree after SCI following a normalized 75 μg dose of phenylephrine (24 ± 18.7% vs 6 ± 11.3%, p=0.007, Figure 2C). Fittingly, to compensate for this larger degree of blood pressure increase, R-R interval AUC over two minutes was also increased after SCI (4245 ± 2217 ms^2^ vs 2471 ± 1703 ms^2^, p=0.02).

**Figure 2:**
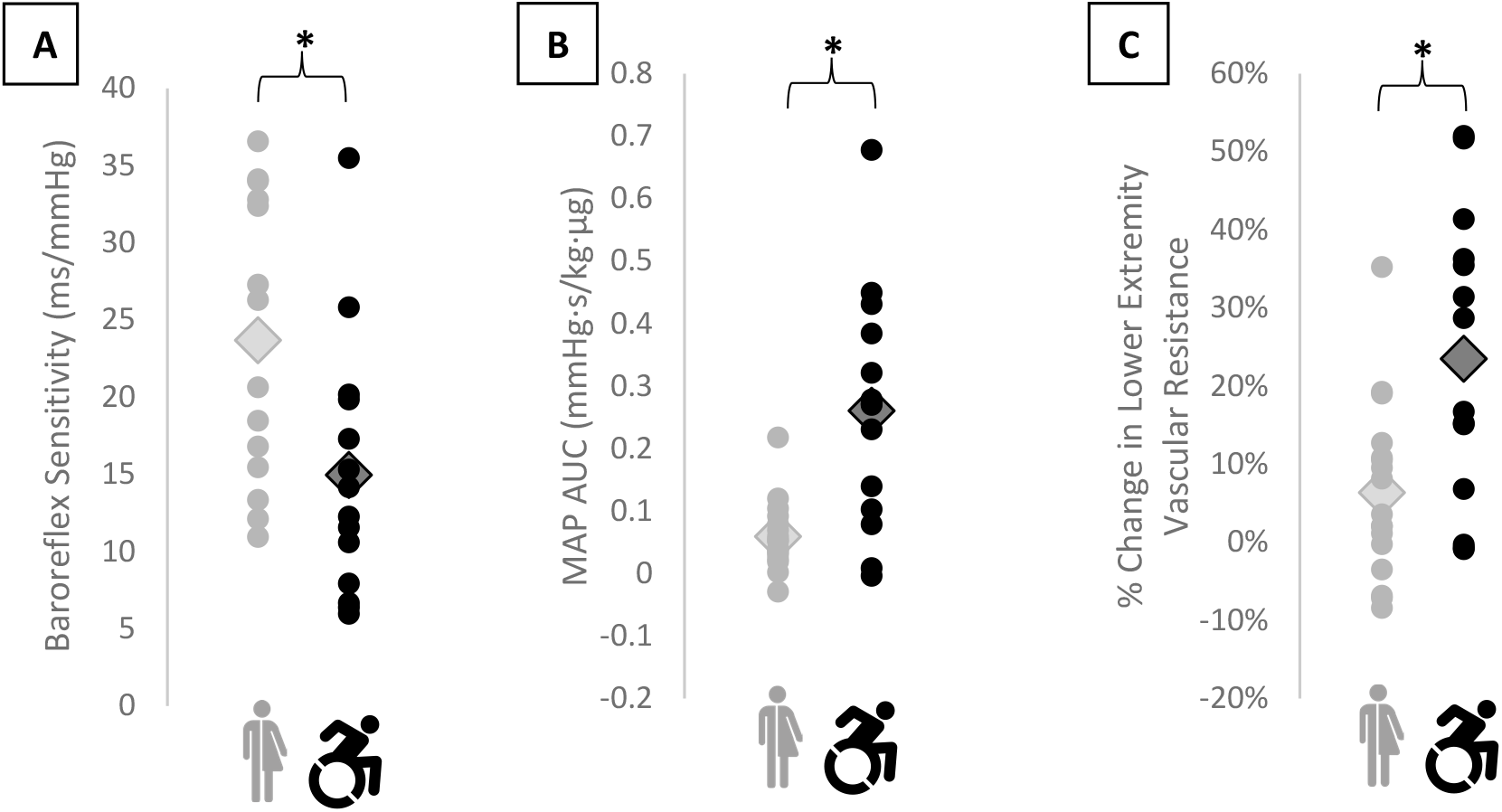
Mean characteristics of the control and SCI cohorts in response to bolus IV phenylephrine. **A)** Baroreflex sensitivity; **B)** 2-minute mean arterial pressure area under the curve following bolus phenylephrine response; **C)** Percentage change in lower extremity vascular resistance from baseline in response to a normalized 75 μg dose, normalized to 75 kg of body weight. *Indicates statistical significance with comparison between control and SCI cohorts. MAP = Mean Arterial Pressure; AUC = Area Under the Curve

For 14 individuals in the SCI cohort, while a discrete range of variables was present, symptoms of AD as measured by the ADFSCI-AD subscore had no meaningful correlation with either baroreflex sensitivity (R^2^=0.09) or MAP AUC (R^2^=0.008) in individuals with SCI (Figure 3).

**Figure 3:**
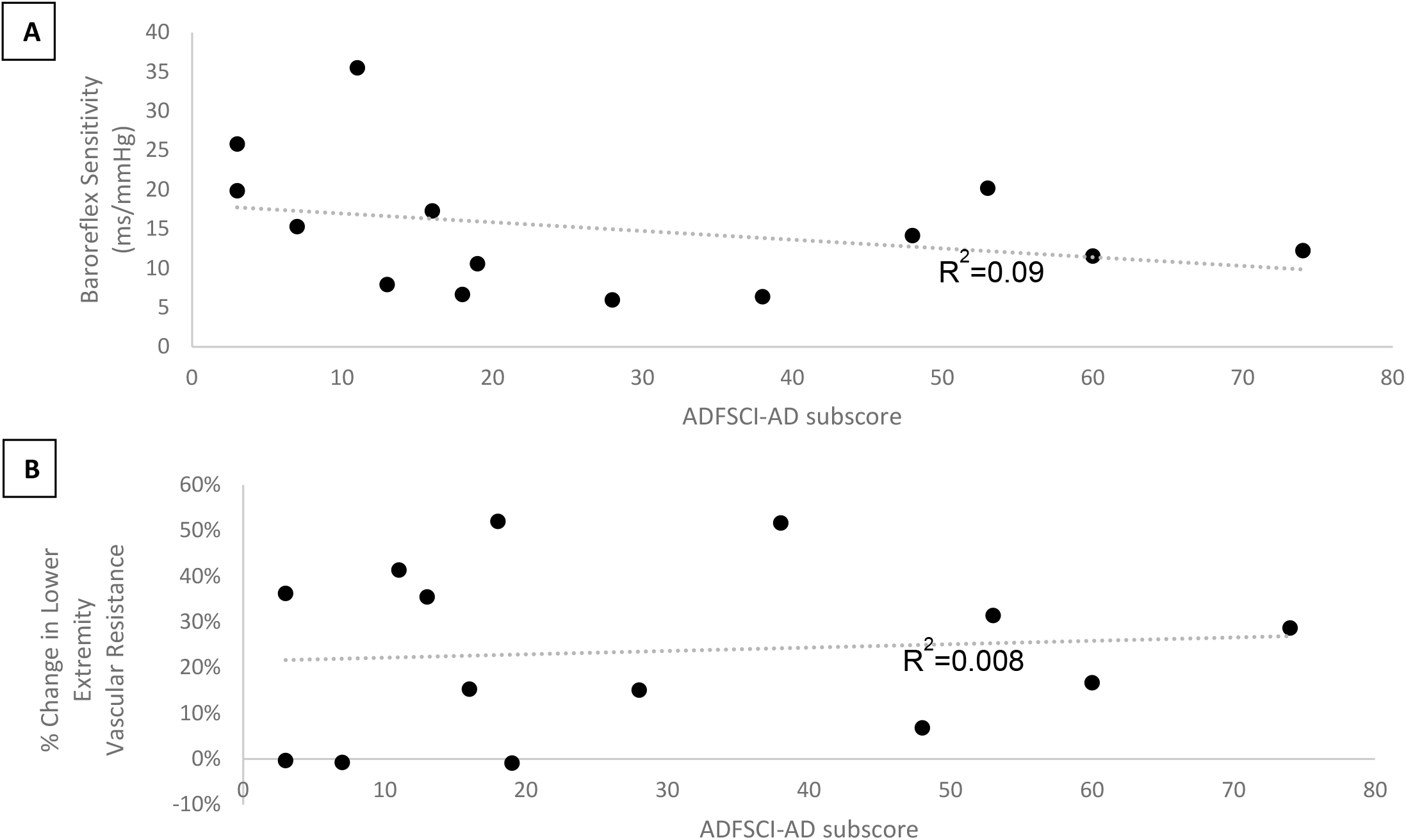
**A**) ADFSCI-AD vs Baroreflex Sensitivity in individuals with SCI; **B)** ADFSCI-AD vs α1 adrenergic receptor sensitivity standardized for 75 kg person in individuals with SCI

## DISCUSSION

On average, individuals with SCI have clearly greater increases in MAP and vascular resistance after IV phenylephrine bolus compared to controls. This is strong evidence of increased α1 adrenergic receptor sensitivity/density (making the vessels hyperresponsive to vasoconstriction). The increase in MAP is likely accentuated further from a decreased ability to buffer these blood pressure increases with compensatory bradycardia (as seen by lower baroreflex sensitivity compared with controls). These findings of increased α1 adrenergic receptor sensitivity and decreased baroreflex sensitivity corroborate past research.^10,15-18^ Importantly, we further demonstrate that while some individuals have altered responses after SCI to these measures, others with SCI have near normal responses. Those with more normal responses may represent individuals with more “autonomically incomplete” SCI, who may have more preserved autonomic regulatory capacity and could, thereby, have less clinical impact from AD. However, deviating from our hypothesis, these variable physiologic measurements of cardiovascular autonomic regulation did not correlate with increased AD symptoms in a meaningful way.

There notably are additional physiologic factors involved in AD pathophysiology (i.e. calcitonin gene related peptide reactive afferent sprouting within the cord, adrenal activation, neural norepinephrine release/uptake, etc.)^26-28^ and it is possible that a more comprehensive model which incorporates these physiologic factors may be needed to better predict AD symptoms. However, practical challenges exist to quantify many of these factors, which are largely accessible only upon tissue biopsy/autopsy.^28,29^

Further potentially clouding our model, symptoms of AD with the ADFSCI-AD survey correlate to the number of AD events individuals with SCI experience, but these objective AD events are not always symptomatic. Thus, the underlying premise that a larger increase in blood pressure with AD (due, for example, to increased α1 adrenergic receptor sensitivity/density) corresponds to more symptoms may need rethinking. Symptoms of AD are no doubt rooted in the underlying physiology, though the experience and perception of burden on quality of life as assessed by the ADFSCI survey may be different between individuals. While this study represents the first known attempt to correlate objective physiologic parameters influencing responses in AD to symptoms individuals experience, symptoms are inherently subjective and more standardized quantification may be needed.

### Limitations

Our study is limited, in part, by the small sample size for both SCI and control groups. We are not able to perform sub-group analysis of different SCI classifications for more nuanced comparisons. Our wide range of NLI further introduces unaccounted variables which future models may be better fit to address.

## CONCLUSION

Individuals with SCI demonstrated both lower baroreflex sensitivity and increased α1 adrenoreceptor sensitivity compared to matched, uninjured controls. These physiologic differences, however, do not correlate with greater past symptom severity of AD.

## Data Availability

All data produced in the present study will be available upon reasonable request to the authors following study completion and analysis.

---

**Supplemental Table 1:** Self-identified racial demographics of study cohorts.

|  | **Controls (n=17)** | **SCI (n=14)** |
| --- | --- | --- |
| Caucasian | 13/17 | 10/14 |
| Asian | 2/17 | 0/14 |
| Hispanic/Latino | 2/17 | 3/14 |
| African-American | 0/17 | 1/14 |

